# Accuracy and usability of saliva and nasal rapid antigen self-testing for detection of SARS-CoV-2 infection in the general population: a head-to-head comparison

**DOI:** 10.1101/2021.12.08.21267452

**Authors:** Ewoud Schuit, Roderick P Venekamp, Irene K Veldhuijzen, Wouter van den Bijllaardt, Suzan D Pas, Joep J J M Stohr, Esther B Lodder, Marloes Hellwich, Richard Molenkamp, Zsofia Igloi, Constantijn Wijers, Irene H Vroom, Carla R S Nagel-Imming, Wanda G H Han, Jan AJW Kluytmans, Susan van den Hof, Janneke H H M van de Wijgert, Karel G M Moons

## Abstract

**Background:** SARS-CoV-2 self-tests may lower the threshold of testing and produce a result quickly. This could support the early detection of infectious cases and reduce further community transmission. However, the diagnostic accuracy of (unsupervised) self-testing with rapid antigen diagnostic tests (Ag-RDTs) is mostly unknown. We therefore conducted a large-scale head-to-head comparison of the diagnostic accuracy of a self-performed SARS-CoV-2 saliva and nasal Ag-RDT, each compared to a molecular reference test, in the general population in the Netherlands.

**Methods:** In this cross-sectional study we consecutively included individuals aged 16 years and older presenting for SARS-CoV-2 testing at three Dutch public health service test sites irrespective of their indication for testing, vaccination status, and symptomatology. Participants were sampled for molecular testing at the test site and received two self-tests (the Hangzhou AllTest saliva self-test and the SD Biosensor nasal self-test by Roche Diagnostics) to perform at home within a few hours without knowledge of their molecular test result. Information on presence and type of symptoms, user experiences, and results of both self-tests were collected via an online questionnaire. For each self-test, sensitivity, specificity, positive and negative predictive values were determined with molecular testing as reference standard.

**Findings:** The SARS-CoV-2 molecular reference test positivity rate was 6.5% in the 2,819 participants. Overall sensitivities with 95% confidence intervals were 46.7% (85/182; 39.3%-54.2%) for the saliva Ag-RDT, and 68.9% (124/180; 61.6%-75.6%) for the nasal Ag-RDT. With a viral load cut-off (≥5.2 log10 SARS-CoV-2 E-gene copies/mL) as a proxy of infectiousness, sensitivities increased to 54.9% (78/142; 46.4%-63.3%) for the saliva Ag-RDT and 83.9% (120/143; 76.9%-89.5%) for the nasal Ag-RDT.

For the nasal Ag-RDT, sensitivities were 78.5% [71.1%-84.8%] and 22.6% [9.6%-41.1%] in those with and without symptoms at the time of sampling, which increased to 90.4% (113/125; 83.8%-94.9%) and 38.9% (7/18; 17.3%-64.3%) after applying the viral load cut-off. In those with and without prior confirmed SARS-CoV-2, sensitivities were 36.8% [19/372; 16.3%-61.6%] and 72.7% [161/2437; 65.1%-79.4%] for the nasal Ag-RDT, which increased to 100% (7/7; 59.0%-100%) and 83.1% (113/126; 75.7%-89.0%) after applying the viral load cut-off.

The diagnostic accuracy of the nasal Ag-RDT did not differ by COVID-19 vaccination status, sex, and age. Specificities were >99%, positive predictive values >70% and negative predictive values >95%, for the saliva Ag-RDT, and >99%, >90%, and >95% for the nasal Ag-RDT, respectively, in most analyses.

Interpreting the results was considered (very) easy for both self-tests.

**Interpretation:** The Hangzhou AllTest self-performed saliva Ag-RDT is not reliable for SARS-CoV-2 infection detection overall nor in the studied subgroups. The SD Biosensor self-performed nasal Ag-RDT had high sensitivity in individuals with symptoms and in those without a prior SARS-CoV-2 infection. The overall accuracy in individuals with symptoms was comparable to that found in previous studies with professional sampling for this Ag-RDT. The extremely low sensitivity of the nasal Ag-RDT in asymptomatic individuals and in individuals who had had a prior SARS-CoV-2 infection is an important finding and warrants further investigation.

**Funding:** Dutch Ministry of Health, Welfare, and Sport.

## Introduction

A molecular test, mainly real-time reverse-transcriptase polymerase chain reaction (RT-PCR), is considered the reference test for SARS-CoV-2 infection detection.^1^ However, molecular tests may take up to 24 hours to deliver a result. Tested individuals are asked to quarantine until they receive a result, which has personal and societal consequences. SARS-CoV-2 rapid antigen diagnostic tests (Ag-RDTs) have shown promising diagnostic accuracies.^2-6^ These Ag-RDTs require no or minimal equipment, provide a result within 15-30 minutes, and can be performed in a range of settings. While Ag-RDTs were initially introduced for use by trained staff at test sites, they can now also be bought in various outlets for self-testing. Such self-testing, without supervision of a trained professional, may potentially lower the threshold to testing and would allow individuals to obtain a test result quickly and at their own convenience. This in turn could support the early detection of infectious cases and reduce community transmission.^7^

Previous studies of self-performed nasal Ag-RDTs showed sensitivities of close to 80% for the Becton Dickinson nasal Ag-RDT in a mixed population of asymptomatic and symptomatic individuals and 82.5% for the SD Biosensor nasal Ag-RDT in symptomatic individuals.^8,9^ Studies employing the SD Biosensor and Abbott Ag-RDTs have shown that supervised nasal self-sampling might be a reliable alternative to nasopharyngeal sampling by a trained professional.^10,11^ However, the sample sizes of these studies were modest, and the nasal self-sampling was supervised instead of self-performed in the home setting. The diagnostic accuracy evidence of self-performed saliva Ag-RDTs is scarce. A recent study found poor performance of four different (unspecified) self-collected saliva Ag-RDTs in a mixed population of symptomatic and asymptomatic individuals, with sensitivities varying from 3.6% to 32.8%, increasing to 5.3% and 41.0% when a “cell culture viability” cut-off was used.^12^ A recent saliva Ag-RDT study employing self-sampling supervised by a trained professional, and testing by that trained professional in a mixed population of symptomatic and asymptomatic individuals found a sensitivity of 66%, which increased to 89% when a cycle threshold (Ct)<30 cut-off was used.^13^

We conducted a large-scale prospective cross-sectional diagnostic accuracy study in the Netherlands of a self-performed saliva and a self-performed nasal Ag-RDT, using a molecular test as the reference standard for each, by head-to-head comparison. We included individuals presenting for routine SARS-CoV-2 testing at Dutch public health service test sites regardless of their reason for testing, vaccination status, and symptomatology at the time of sampling. A secondary aim was to evaluate user experiences and preferences for both self-performed Ag-RDTs.

## Methods

The study is reported according to the STARD 2015 guidelines: an updated list of essential items for reporting diagnostic accuracy studies.^14^

### Study design and population

This large prospective cross-sectional diagnostic test accuracy study was embedded within the Dutch public testing infrastructure. Public testing in the Netherlands, by default molecular testing, is free-of-charge but only available for government-approved test indications. At the time of the study (9 to 26 September 2021), testing indications included having symptoms of a potential SARS-CoV-2 infection; having been identified as a close contact of a SARS-CoV-2 index case via traditional contact-tracing or the contact-tracing app regardless of symptomatology at the time of notification; having tested positive on a nasal self-test performed outside of this study; or having returned from a country on the government’s list of high risk countries.^15^ Participants were recruited consecutively at three Dutch public health service COVID-19 test sites, located in West-Brabant (Roosendaal), Central- and Northeast Brabant (Tilburg), and Rotterdam-Rijnmond (Zuidland). Individuals were considered eligible if they were aged 16 years or older, and willing and able to sign a digital informed consent in Dutch.

The study was conducted when the SARS-CoV-2 prevalence was 8.2% with the Delta variant as the dominant variant in the Netherlands (99.9%) during the entire study period.^16-18^

### Inclusion procedure

Individuals who attended one of the participating test sites for a routine molecular SARS-CoV-2 test were asked by the test site staff whether they were willing to participate. If interested, they received a participant information letter, the saliva and the nasal Ag-RDT together with an instruction manual, and an email with a study participation link to access study documentation. Next, trained test site staff took a swab for routine molecular testing (see below). Participants were asked to provide informed consent electronically via the participation link after arriving home, to subsequently perform both self-tests as soon as possible but within three hours (the saliva test first, followed by the nasal test), and to complete a short online baseline questionnaire. This included questions on demographics; presence, type, and onset of COVID-19-related symptoms; indication for testing; vaccination status including type of vaccine and vaccination date(s); the results of the two Ag-RDTs that they had just performed; and their user experiences with and opinions about both Ag-RDTs (supplementary material 1). Participants whose online questionnaire was not completed within three hours of their test site visit were contacted by a call center with the request to perform both self-tests and complete the questionnaire as soon as possible.

Ten days after their test site visit, participants received an email asking them to complete an online follow-up questionnaire. The follow-up questionnaire included questions on COVID-19-related symptoms and SARS-CoV-2 testing during follow-up (supplementary material 2) to capture any infections that may have been missed by the baseline molecular test.

### Specimen collection and testing

Molecular reference test sampling was performed by trained test site staff. While molecular testing was always used as the reference standard, the three test sites used slightly different sampling methods and the three affiliated centralized laboratories used slightly different molecular testing methods (supplementary material 3). Briefly, the Roosendaal site combined oropharyngeal-nasal sampling with RT-PCR testing on a Roche cobas 8800 platform. The Tilburg site combined oropharyngeal and nasopharyngeal sampling with the Abbott Alinity M SARS-CoV-2 assay or in-house RT-PCR19; samples that tested positive by RT-PCR in Tilburg were subsequently tested on the Roche cobas 8800 platform in Roosendaal to obtain Ct values for viral load calculation. The Zuidland site combined oropharyngeal and nasopharyngeal sampling with RT-PCR on a Roche cobas 6800 platform.

Participants performed the saliva and nasal Ag-RDTs themselves in their own homes (i.e., unsupervised) according to the instructions in the manual that they received at the test site; those instructions were identical to those provided by the manufacturers but translated into Dutch. The saliva Ag-RDT that we used was the COVID-19 Antigen Rapid Test (Oral Fluid) for Self-testing by Hangzhou AllTest Biotech Co. Ltd., and is a so-called spitting test: participants had to spit in a funnel connected to a tube. The nasal Ag-RDT that we used was the SD Biosensor SARS-CoV-2 Rapid Antigen Test Nasal for self-testing, distributed by Roche Diagnostics (supplementary material 3). Both tests are CE-marked, and the Hangzhou AllTest was the only saliva self-test with CE-marking at the time of study conception. Participants interpreted their Ag-RDT test results visually in accordance with the instructions and this interpretation was always done before they had received their molecular test result. Vice versa, the Ag-RDTs results were not available to those in the study team assessing the molecular test results. Participants received their molecular test results from the public health services test site that they attended conform routine practice to direct any further management (such as quarantine advice, if applicable).

### Outcomes and statistical analyses

The primary outcomes were the diagnostic accuracies (sensitivity, specificity, positive and negative predictive values with corresponding 95% confidence intervals [CI]) of each self-test, with molecular testing as the reference standard. We performed a complete cases analysis because the number of individuals without molecular test or Ag-RDT results was very low (n=147 (5.0%) for the saliva Ag-RDT, and n=131 (4.4%) for the nasal Ag-RDT); Figure 1).

**Figure 1.**
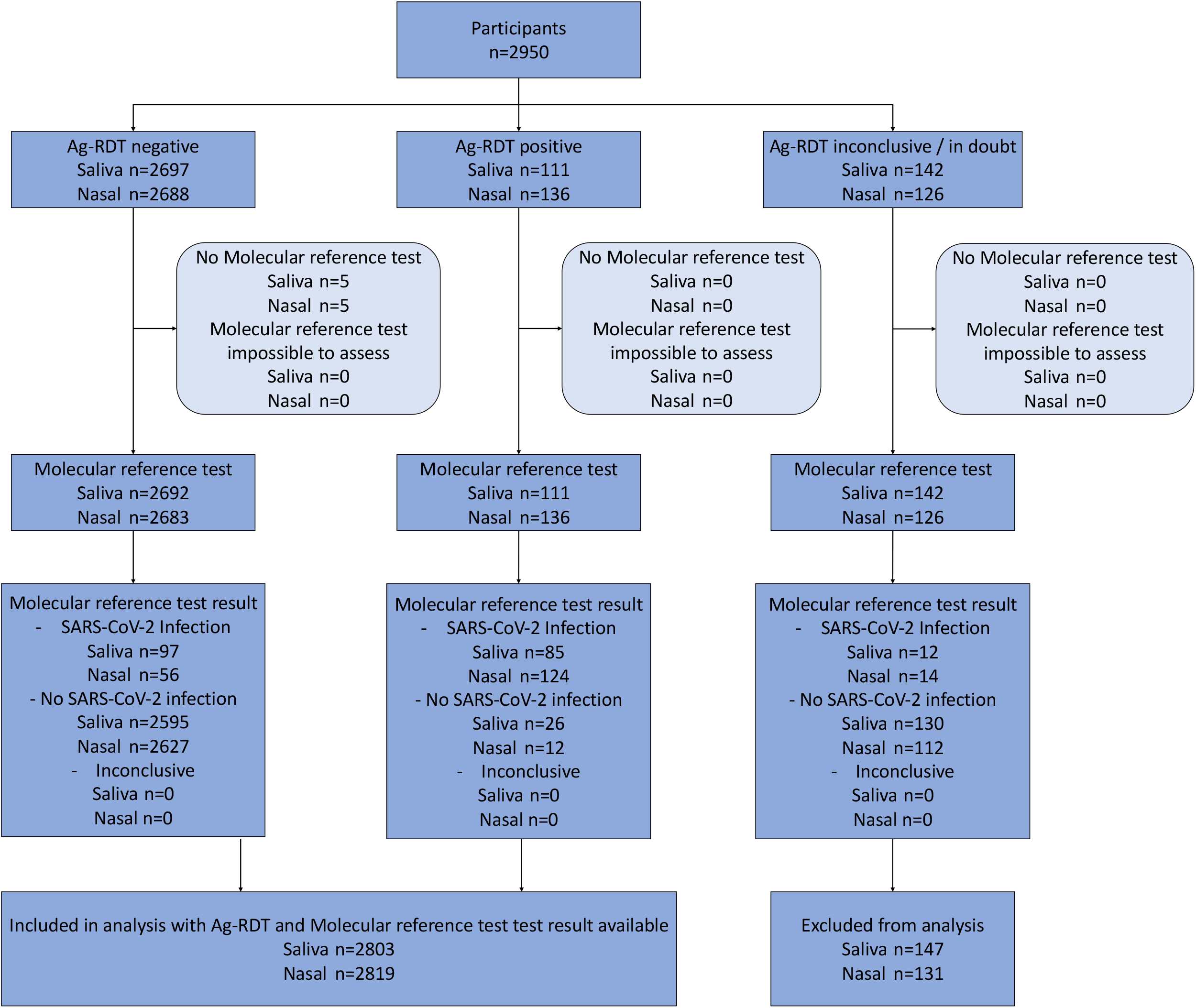
Flow of study participants. Saliva = COVID-19 Antigen Rapid Test (Oral Fluid) for Self-testing by Hangzhou AllTest Biotech Co., Ltd., Nasal = SD Biosensor SARS-CoV-2 Rapid antigen test Nasal for self-testing by Roche Diagnostics.

Secondary outcomes were diagnostic accuracies stratified by presence of symptoms at the time of sampling (yes or no), COVID-19 vaccination status (vaccinated with at least one dose yes or no), having had a prior SARS-CoV-2 infection (yes or no), gender (female or male), and age (≥16 to ≤40 or >40 to ≤65 or >65 years). Additional secondary outcomes included all of the above but after using a viral load cut-off as a proxy of infectiousness (≥5.2 log10 SARS-CoV-2 E-gene copies/mL), which was the viral load cut-off above which 95% of people with a positive molecular test had a positive virus culture in a recent study by our group3. In addition, we assessed the user experiences and preferences for both self-performed Ag-RDTs.

Finally, using the follow-up questionnaire, we determined whether participants who received a negative molecular test result at baseline had tested positive in the subsequent 10 days by either molecular test or Ag-RDT.

### Sample size calculation

Previous nasal Ag-RDTs performance studies in symptomatic individuals found sensitivities around 85% when performed by trained staff2 and around 80% when used as a self-test.^8^ A recent accuracy study in The Netherlands that quantified the accuracy of saliva Ag-RDT performed by trained professionals, found an overall sensitivity of 66% and around 89% when using a Ct<30 cut-off.^13^ We therefore based our sample size calculation on an expected sensitivity of 80% for each self-performed Ag-RDT, with a margin of error of 7%, type I error of 5% and power of 80%. Hence, we aimed for approximately 140 positive molecular reference tests per Ag-RDT evaluation. When planning the study (June 2021), we anticipated a SARS-CoV-2 prevalence (based on molecular test positivity rates) in our target population of around 5%, and closely monitored molecular test positivity rates over time to prolong recruitment as needed.

## Results

Between 9 and 26 September 2021, 2,950 individuals participated in the study (Figure 1). An Ag-RDT result with matching molecular reference test result were available for 2,803 saliva Ag-RDT users (95.0%) and 2,819 nasal Ag-RDT users (95.5%).

The demographic characteristics of study participants are presented in Table 1 and Supplementary Table 1. The mean age was 41 years (standard deviation 15.5) and 61% was female. The majority was vaccinated (85% once; 74% twice), had not had a previous SARS-CoV-2 infection (87%), and was symptomatic at the time of sampling (83%). These characteristics were comparable across test sites, although participants presenting at the Zuidland location were less often vaccinated than those presenting in Roosendaal and Tilburg (76.8% vs. 85.2% vs. 87.5%). Table 2 and Figure 2 show the results of the primary analysis and the secondary stratified analyses. Supplementary Figure 1 presents them after the application of a viral load cut-off as a proxy for infectiousness. Supplementary Tables S1 and S2 show 2×2 tables for both Ag-RDTs. The main findings are presented in the text below.

**Table 1.**
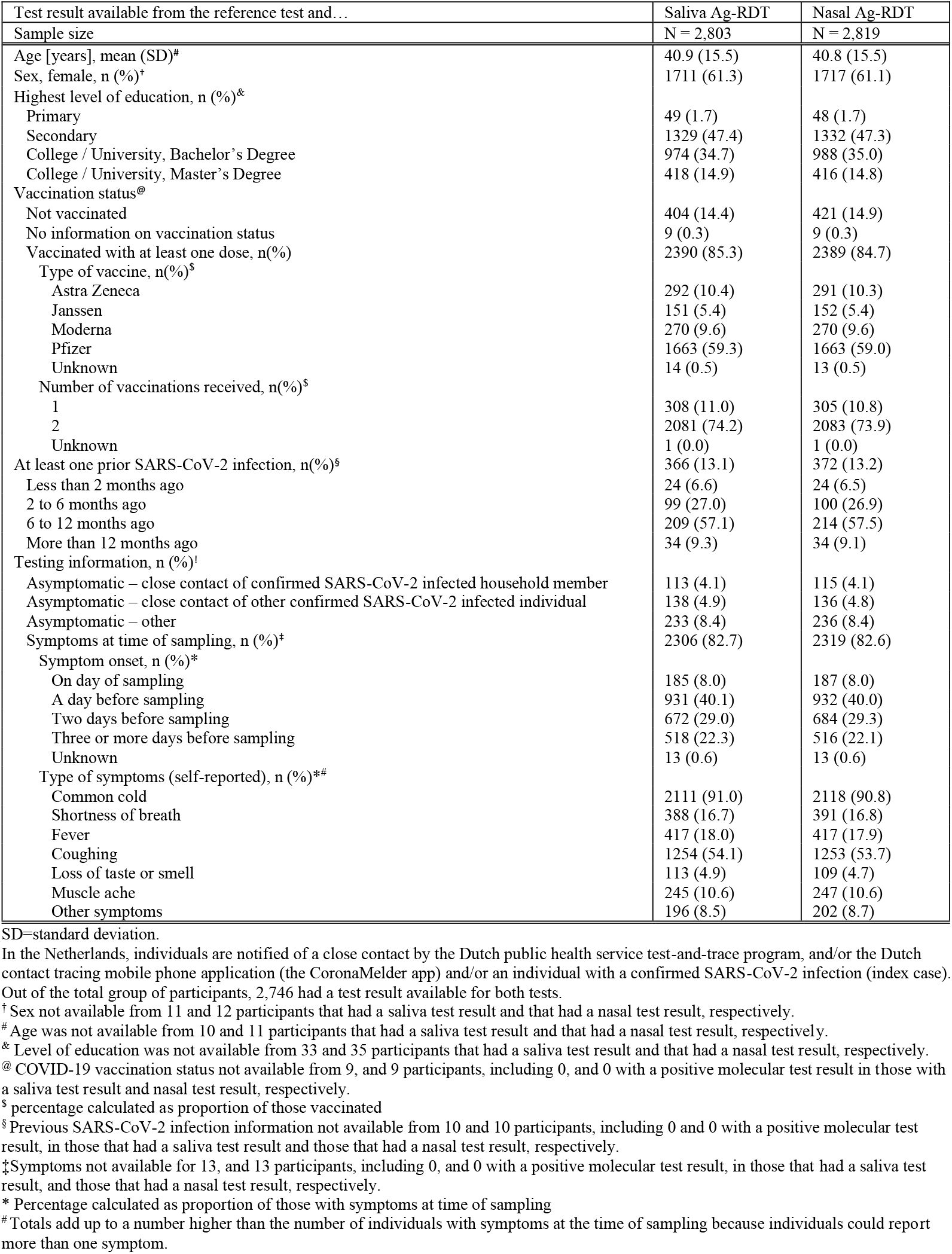
Baseline characteristics of the study population, stratified by type of rapid antigen test.

**Table 2.** Diagnostic accuracy parameters for the saliva and nasal Ag-RDTs. Values are percentages (95% confidence interval) unless stated otherwise.

| Analysis | No. | Prevalence*<br>[%] | Sensitivity<br>(95%CI)<br>[%] | Specificity<br>(95%CI)<br>[%] | PPV<br>(95%CI)<br>[%] | NPV [%] (95%CI) |
| --- | --- | --- | --- | --- | --- | --- |
| **COVID-19 Antigen Rapid Test (Oral Fluid) for Self-testing (Hangzhou AllTest Biotech Co., Ltd.)** |  |  |  |  |  |  |
| **Primary analysis** | 2,803 | 6.5 | 46.7 (39.3 to 54.2) | 99.0 (98.5 to 99.4) | 76.6 (67.6 to 84.1) | 96.4 (95.6 to 97.1) |
| **Secondary (stratified) analysis:** |  |  |  |  |  |  |
| Infectiousness viral load cut-off¶ | 2,788 | 5.1 | 54.9 (46.4 to 63.3) | 98.8 (98.3 to 99.2) | 70.9 (61.5 to 79.2) | 97.6 (97.0 to 98.2) |
| Symptoms present at sampling‡: |  |  |  |  |  |  |
| Yes | 2,306 | 6.5 | 51.0 (42.7 to 59.2) | 98.8 (98.3 to 99.2) | 75.5 (66.0 to 83.5) | 96.6 (95.8 to 97.4) |
| No | 484 | 6.4 | 25.8 (11.9 to 44.6) | 99.8 (98.8 to 100) | 88.9 (51.8 to 99.7) | 95.2 (92.8 to 96.9) |
| Vaccinated (at least one)®: |  |  |  |  |  |  |
| Yes | 2,390 | 4.8 | 47.0 (37.6 to 56.5) | 99.0 (98.5 to 99.4) | 71.1 (59.5 to 80.9) | 97.4 (96.6 to 98.0) |
| No | 404 | 16.6 | 46.3 (34.0 to 58.9) | 98.8 (97.0 to 99.7) | 88.6 (73.3 to 96.8) | 90.2 (86.7 to 93.1) |
| Previous SARS-CoV-2 infection§: |  |  |  |  |  |  |
| Yes | 366 | 5.5 | 20.0 (5.7 to 43.7) | 99.1 (97.5 to 99.8) | 57.1 (18.4 to 90.1) | 95.5 (92.9 to 97.4) |
| No | 2,427 | 6.7 | 50.0 (42.1 to 57.9) | 99.0 (98.5 to 99.4) | 77.9 (68.7 to 85.4) | 96.5 (95.7 to 97.2) |
| Sex†: |  |  |  |  |  |  |
| Female | 1,711 | 6.3 | 38.0 (28.8 to 47.8) | 99.2 (98.6 to 99.6) | 75.9 (62.4 to 86.5) | 96.0 (94.9 to 96.9) |
| Male | 1,081 | 6.8 | 60.3 (48.1 to 71.5) | 98.7 (97.8 to 99.3) | 77.2 (64.2 to 87.3) | 97.2 (96.0 to 98.1) |
| Age [years]#: |  |  |  |  |  |  |
| ≥16 to ≤40 | 1,450 | 6.3 | 41.3 (31.1 to 52.1) | 98.9 (98.2 to 99.4) | 71.7 (57.7 to 83.2) | 96.1 (95.0 to 97.1) |
| >40 to ≤65 | 1,135 | 6.3 | 50.0 (38.0 to 62.0) | 99.2 (98.4 to 99.6) | 80.0 (65.4 to 90.4) | 96.7 (95.5 to 97.7) |
| >65 | 208 | 8.2 | 64.7 (38.3 to 85.8) | 99.0 (96.3 to 99.9) | 84.6 (54.6 to 98.1) | 96.9 (93.4 to 98.9) |
| **SD Biosensor SARS-CoV-2 Rapid antigen test Nasal by Roche Diagnostics ('Nasal'),** |  |  |  |  |  |  |
| **Primary analysis** | 2,819 | 6.4 | 68.9 (61.6 to 75.6) | 99.5 (99.2 to 99.8) | 91.2 (85.1 to 95.4) | 97.9 (97.3 to 98.4) |
| **Secondary (stratified) analysis:** |  |  |  |  |  |  |
| Infectiousness viral load cut-off¶ | 2,804 | 5.1 | 83.9 (76.9 to 89.5) | 99.5 (99.2 to 99.7) | 90.2 (83.9 to 94.7) | 99.1 (98.7 to 99.5) |
| Symptoms present at sampling‡: |  |  |  |  |  |  |
| Yes | 2,319 | 6.4 | 78.5 (71.1 to 84.8) | 99.5 (99.2 to 99.8) | 92.1 (86.0 to 96.2) | 98.5 (97.9 to 99.0) |
| No | 487 | 6.4 | 22.6 (9.6 to 41.1) | 99.6 (98.4 to 99.9) | 77.8 (40.0 to 97.2) | 95.0 (92.6 to 96.8) |
| Vaccinated (at least one)®: |  |  |  |  |  |  |
| Yes | 2,389 | 4.5 | 70.1 (60.5 to 78.6) | 99.6 (99.2 to 99.8) | 88.2 (79.4 to 94.2) | 98.6 (98.0 to 99.0) |
| No | 421 | 17.3 | 67.1 (55.1 to 77.7) | 99.4 (97.9 to 99.9) | 96.1 (86.5 to 99.5) | 93.5 (90.5 to 95.8) |
| Previous SARS-CoV-2 infection§: |  |  |  |  |  |  |
| Yes | 372 | 5.1 | 36.8 (16.3 to 61.6) | 99.2 (97.5 to 99.8) | 70.0 (34.8 to 93.3) | 96.7 (94.3 to 98.3) |
| No | 2,437 | 6.6 | 72.7 (65.1 to 79.4) | 99.6 (99.3 to 99.8) | 92.9 (86.9 to 96.7) | 98.1 (97.5 to 98.6) |
| Sex†: |  |  |  |  |  |  |
| Female | 1,717 | 6.2 | 68.9 (59.1 to 77.5) | 99.6 (99.1 to 99.8) | 91.2 (82.8 to 96.4) | 98.0 (97.2 to 98.6) |
| Male | 1,090 | 6.7 | 69.9 (58.0 to 80.1) | 99.5 (98.9 to 99.8) | 91.1 (80.4 to 97.0) | 97.9 (96.8 to 98.7) |
| Age [years]#: |  |  |  |  |  |  |
| ≥16 to ≤40 | 1,463 | 6.4 | 66.7 (56.1 to 76.1) | 99.5 (99.0 to 99.8) | 89.9 (80.2 to 95.8) | 97.8 (96.9 to 98.5) |
| >40 to ≤65 | 1,139 | 6.1 | 72.9 (60.9 to 82.8) | 99.6 (99.0 to 99.9) | 92.7 (82.4 to 98.0) | 98.2 (97.3 to 98.9) |
| >65 | 206 | 7.8 | 68.8 (41.3 to 89.0) | 99.5 (97.1 to 100) | 91.7 (61.5 to 99.8) | 97.4 (94.1 to 99.2) |

PPV=positive predictive value; NPV=negative predictive value.
\*SARS-CoV-2 infection based on molecular test result.
¶Viral load cut-off for infectiousness, defined as viral load above which 95% of people with a positive RT-PCR test result had a positive viral culture<sup>6</sup>, was 5.2 log<sub>10</sub> SARS-CoV-2 E-gene copies/mL.
§Viral load unavailable for 19 individuals that had a saliva test result and 19 individuals that had a nasal test result.
‡Symptoms not available for 13, and 13 participants, including 0, and 0 with a positive molecular test result, in those that had a saliva test result, and those that had a nasal test result, respectively.
® COVID-19 vaccination status not available from 9, and 9 participants, including 0, and 0 with a positive molecular test result in those with a saliva test result and nasal test result, respectively.
§ Previous SARS-CoV-2 infection information not available from 10 and 10 participants, including 0 and 0 with a positive molecular test result, in those that had a saliva test result and those that had a nasal test result, respectively.
† Sex not available from 11 and 12 participants that had a saliva test result and that had a nasal test result, respectively.

**Figure 2.**
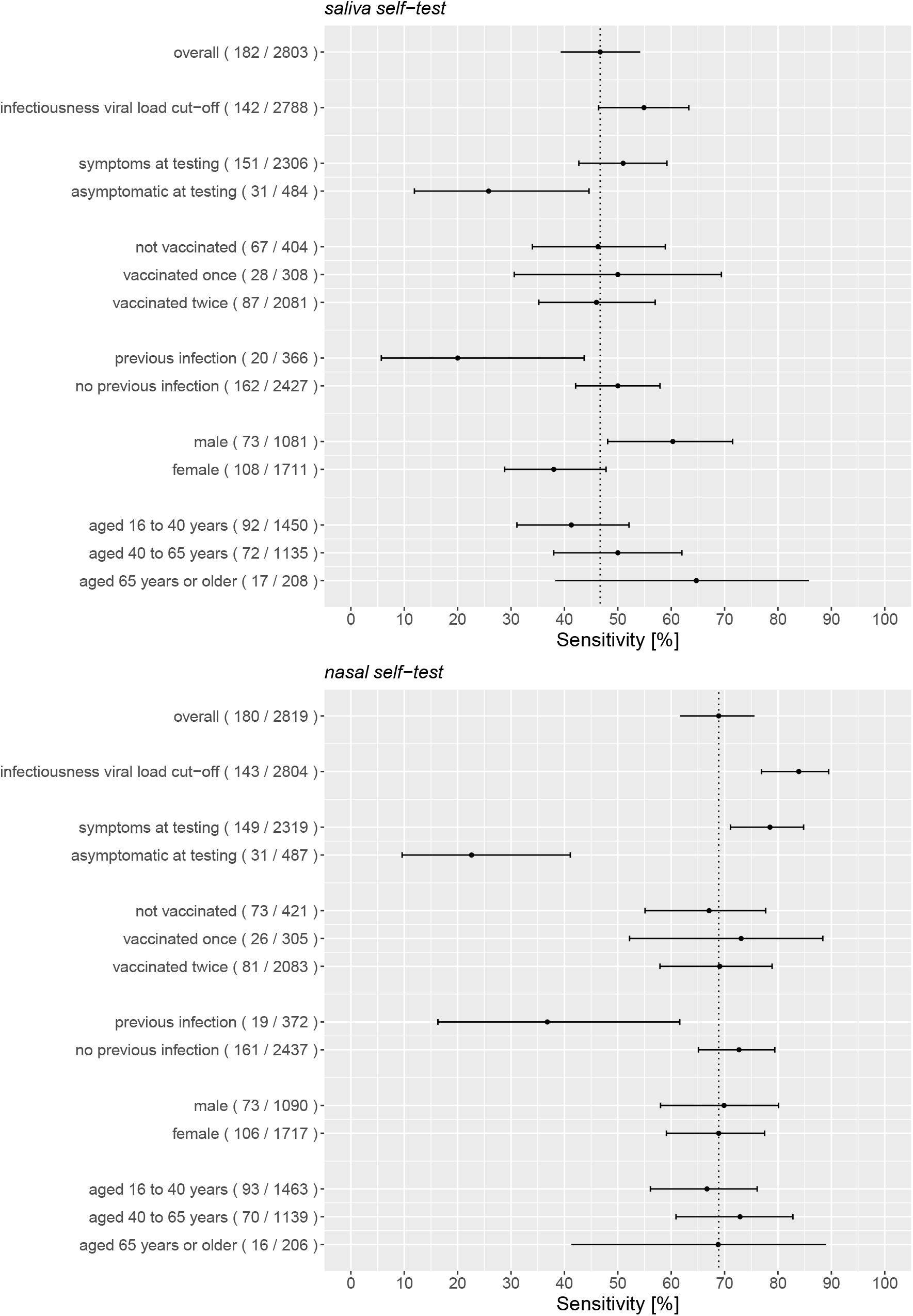
Sensitivities with 95% confidence intervals of the antigen rapid test-molecular reference standard test comparisons stratified according to symptomatology, COVID-19 vaccination status, previous infection status, sex, and age. The vertical line indicates the sensitivity of the Ag-RDT in the overall study population, and the number of positive molecular tests out of the total or subgroup between parentheses.

### Self-performed saliva Ag-RDT

#### Overall test accuracy

SARS-CoV-2 molecular test positivity was 6.5% (182/2803) and overall sensitivity was 46.7% (85/182; 95% CI 39.3%-54.2%; Table 2, Figure 2). Among those with a positive molecular test result, the percentage of participants with a viral load above the cut-off as a proxy for infectiousness was 78.0% (142/182). Using this viral load cut-off, the overall sensitivity was 54.9% (78/142; 46.4%-63.3%; Table 2, Figure 2). Specificities were around 99% in both analyses; positive and negative predictive values are presented in Table 2.

#### Stratified analyses

The sensitivity was 51.0% (77/151; 42.7%-59.2%) in participants who were symptomatic at the time of testing and 25.8% (8/31; 11.9%-44.6%) in participants who were asymptomatic (Table 2, Figure 2). After application of a viral load cut-off as a proxy for infectiousness, these sensitivities were 57.3% (71/124; 48.1%-66.1%) and 38.9% (7/18; 17.3%-64.3%), respectively (Supplementary Figure 1). The sensitivity was 20.0% (4/20; (5.7%-43.7%) in participants who had had a previous SARS-CoV-2 infection and 50.0% (81/162; 42.1%-57.9%) in participants who had never had a prior infection (Table 2, Figure 2). After applying the viral load cut-off, these sensitivities were 50.0% (4/8; 15.7%-84.3%) and 55.2% (74/134; 46.4%-63.8%), respectively (Supplementary Figure 1). In males the sensitivity was higher than in females, 60.3% (44/73; 48.1%-71.5%) versus 38.0% (41/108; 28.8%-47.8%), and increased slightly by increasing age (Table 2, Figure 2). We found no evidence of a differential impact on diagnostic accuracy by COVID-19 vaccination status. The sensitivities after application of a viral load cut-off as a proxy for infectiousness, are presented in Supplementary Figure 1. Specificities were >99%, and positive predictive values >70% and negative predictive values >95% in most analyses (Table 2).

### Self-performed nasal Ag-RDT

#### Overall test accuracy

SARS-CoV-2 molecular test positivity was 6.4% (180/2819) and overall sensitivity was 68.9% (124/180; 61.6%-75.6%; Table 2, Figure 2). Among those with a positive molecular test result, the percentage of participants with a viral load above the cut-off as a proxy for infectiousness was 79.4% (143/180). Using this viral load cut-off, the overall sensitivity was 83.9% (120/143; 76.9%-89.5%; Table 2, Figure 2). Specificities were 99.5% in both analyses; positive and negative predictive values are presented in Table 2.

#### Stratified analyses

The sensitivity was 78.5% (117/149; 71.1%-84.8%) in participants who were symptomatic at the time of testing and 22.6% (7/31; 9.6%-41.1%) in participants who were asymptomatic (Table 2, Figure 2). After application of a viral load cut-off as a proxy for infectiousness, these sensitivities were 90.4% (113/125; 83.8%-94.9%) and 38.9% (7/18; 17.3%-64.3%), respectively (Supplementary Figure 1). On average, the viral load was lower in asymptomatic than in symptomatic individuals with a positive molecular reference test (Supplementary Figure 2). The sensitivity was 36.8% (7/19; 16.3%-61.6%) in participants who had had a previous SARS-CoV-2 infection and 72.7% (117/161; 65.1%-79.4%) in participants who had never had a prior infection. After applying the viral load cut-off, these sensitivities were 100.0% (7/7; 59.0%-100.0%) and 83.1% (113/126; 75.7%-89.0%), respectively (Supplementary Figure 1). We found no evidence of a differential impact on diagnostic accuracy by COVID-19 vaccination status, sex, and age (Figure 2, supplementary Table S3). Specificities were >99%, and positive predictive values >90% and negative predictive values >95% in most analyses (Table 2).

Diagnostic test accuracy results for both Ag-RDTs were the same when the analysis populations were limited to the 2,746 participants for whom all three test results were available (data not shown).

### User experiences and preferences

A larger proportion of participants indicated that taking a sample was easy or very easy for the self-performed saliva Ag-RDT than for the self-performed nasal Ag-RDT (80.4% vs. 66.8%). Most participants reported that reading the test result was easy or very easy for both tests (92.2% vs. 92.4%; Table 3). After performing both Ag-RDTs, but before receiving the molecular test result, 55.6% of the participants reported to prefer the saliva over the nasal Ag-RDT, 9.3% reported to prefer the nasal Ag-RDT, and 32.8% indicated to have no preference.

**Table 3.** Usability of the rapid antigen saliva and nasal Ag-RDTs.

| N = 2,950 | Saliva Ag-RDT | Nasal Ag-RDT |
| --- | --- | --- |
| Difficulty taking a sample |  |  |
| Very easy | 1286 (43.6) | 726 (24.6) |
| Easy | 1087 (36.8) | 1245 (42.2) |
| Medium | 380 (12.9) | 613 (20.8) |
| Hard | 117 (4.0) | 265 (9.0) |
| Very hard | 16 (0.5) | 35 (1.2) |
| Unknown | 64 (2.2) | 66 (2.2) |
| Difficulty reading the test result |  |  |
| Very easy | 1997 (67.7) | 1940 (65.8) |
| Easy | 724 (24.5) | 785 (26.6) |
| Medium | 110 (3.7) | 128 (4.3) |
| Hard | 39 (1.3) | 23 (0.8) |
| Very hard | 15 (0.5) | 9 (0.3) |
| Unknown | 65 (2.2) | 65 (2.2) |
| The user manual was clear |  |  |
| Agree | 2533 (85.9) | n/a |
| In doubt | 266 (9.0) | n/a |
| Disagree | 86 (2.9) | n/a |
| Unknown | 65 (2.2) | n/a |
| The test went well |  |  |
| Agree | 2514 (85.2) | n/a |
| In doubt | 322 (10.9) | n/a |
| Disagree | 50 (1.7) | n/a |
| Unknown | 65 (2.2) | n/a |
| If I could, I would use the test at home |  |  |
| Agree | 2554 (86.6) | n/a |
| In doubt | 89 (3.0) | n/a |
| Disagree | 239 (8.1) | n/a |
| Unknown | 68 (2.3) | n/a |
| Do you prefer one test over the other |  |  |
| No | 969 (32.8) |  |
| Yes, saliva Ag-RDT preferred | 1639 (55.6) |  |
| Yes, nasal Ag-RDT preferred | 275 (9.3) |  |
| Unknown | 67 (2.3) |  |

n/a = not available

### Follow-up

Follow-up information was available for 72% of participants (Table S3), of whom 1994 participants had a negative molecular test result at baseline. Of the latter group, 249/1994 (12.5%) were re-tested within 10 days, and 7/249 (2.8%) tested positive, wherein we did not know whether it was a new infection, or the initial molecular test was false negative.

## Discussion

This largest diagnostic accuracy evaluation of two unsupervised self-performed Ag-RDTs to date showed a low overall sensitivity (46.7%) of the Hangzhou AllTest Biotech saliva self-test. Applying a viral load cut-off as a proxy for infectiousness did not improve the overall sensitivity meaningfully (54.9%) nor in any of the studied subgroups, with all sensitivities remaining far below the WHO standard of 80%^20^.

The study showed better performance of the SD Biosensor nasal self-test with an overall sensitivity of 68.9%, increasing to 83.9% when the viral load cut-off was applied. The sensitivities were much higher in the 2,319 symptomatic participants than in the 487 asymptomatic participants (78.5% and 22.6%, respectively), and in the 2,417 individuals who never had COVID-19 in the past compared to the 372 individuals who did (72.7% and 36.8%, respectively), reaching sensitivities (with sufficient precision) above the WHO-recommended 80% in symptomatic individuals and in individuals without a previous infection after applying the viral load cut-off. The sensitivities in asymptomatic individuals and in individuals who had had COVID-19 in the past have wider 95% confidence intervals and should therefore be interpreted with caution. We recommend additional research in those groups. The diagnostic accuracy of this nasal self-test did not differ by COVID-19 vaccination status, sex, and age.

### Discussion of the saliva self-test results

We identified two previous studies in the scientific literature; the sensitivities observed in our study were in between those found in the two studies. A Czech study evaluated four saliva Ag-RDTs and found sensitivities of 15% for a saliva test that required spitting in a cup, and 3.6%, 25.5%, and 32.8% for saliva tests requiring sucking on a sponge, in comparison with RT-PCR.^12^ These sensitivities improved slightly after using a “cell culture viability” cut-off but remained well below 50%. The sampling was done by the participants themselves but supervised by trained personnel. Samples sizes were modest, ranging from 98 to 407 participants per evaluated test. A recent Dutch study of the SD Biosensor saliva test with 789 participants found a sensitivity of 66.1%, increasing to 88.6% when Ct<30, and to 96.7% when viral culturability, were used as cut-offs.^13^ In the Dutch study, saliva was collected by letting nasal and cough discharge drool into a collection device and was supervised by trained test site staff. Sensitivity was lower (60%) in asymptomatic participants but only 10 asymptomatic participants tested RT-PCR positive. We tested the analytical performance of the Hangzhou and SD Biosensor lateral flow test devices on calibrated samples and found that both test devices performed (equally) well (Supplement 3). We therefore hypothesize that the widely ranging sensitivity results for saliva Ag-RDTs may be due to high variability in saliva sampling methods (spitting vs. sucking vs. drooling) and/or high variability in the quantity and quality of sample self-obtained by different individuals. Furthermore, saliva specimens may on average contain lower SARS-CoV-2 viral loads than upper respiratory tract samples. Studies have shown that saliva viral loads are usually sufficiently high for detection by molecular methods^21-23^, but they may not be sufficiently high for detection by self-performed Ag-RDTs.

We saw trends of reduced diagnostic accuracy in persons without symptoms or with previous SARS-CoV-2 infection. These trends were like the trends that we observed for the nasal self-test and are discussed below. We also saw trends by gender and age, with sensitivities for men and for persons aged over 65 reaching around 60%, which is still well below the WHO-recommended 80%.^20^ The saliva Ag-RDT evaluation studies to date did not stratify by gender and age^12,13^, and the nasal Ag-RDT studies, including the nasal self-test that we evaluated in this study, did not show these trends.^3,6^ We recommend that future saliva Ag-RDT evaluations stratify by gender and age to investigate this further.

## Discussion of the nasal self-test results

The diagnostic performance of Ag-RDTs combined with nasopharyngeal sampling done by trained personnel or by individuals themselves have been evaluated extensively by us and others.^2-6,8,9,12^ The above-mentioned Czech study also evaluated an Ag-RDT in combination with anterior nasal sampling done by trained personnel.^12^ These studies found good performance (70-80%) with nasopharyngeal sampling, but lower performance with anterior nasal sampling (45-55%), and also lower performance in asymptomatic individuals (50-60%), regardless of self or professional sampling. Our study showed that nasal self-sampling with the SD Biosensor Ag-RDT provided good sensitivity, which equaled the sensitivity of Ag-RDTs found in other studies in which the nasal sampling was done by a trained professional^10,11^, but only for individuals who have symptoms at the time of testing. We found a very low sensitivity of this self-performed nasal Ag-RDT of only 23% in asymptomatic individuals, which is much lower than the sensitivities found in our previous studies using nasopharyngeal or oropharyngeal combined with nasal sampling done by trained personnel.^3,6^ This difference in performance persisted after applying a viral load cut-off. It is currently unclear why the sensitivities of the nasal Ag-RDT self-test differed depending on the presence of symptoms, even after applying the viral load cut-off. We tested the analytical performance of the SD Biosensor lateral flow test device on calibrated samples and found that the test device itself performed well (Supplement 3). We hypothesize that the difference in sensitivity may be explained by the difference in viral load distributions in asymptomatic and symptomatic individuals. In addition, it may be more difficult for asymptomatic individuals (i.e., with a dry nose) to retrieve sufficient nasal fluid by self-swabbing. The former hypothesis is supported by the fact that the sensitivity of the saliva self-test was also lower in asymptomatic than symptomatic individuals, but both hypotheses might play a role.

We also found a low sensitivity (36.8%) of the nasal self-test in individuals who had had COVID-19 in the past. These results should be interpreted with caution due to the small group sizes: only 19 participants with a positive molecular test reported having had COVID-19 (16 of whom were symptomatic at the time of testing), and only seven of them had a viral load above the viral load cut-off (six of whom were symptomatic). However, similar trends were observed for the saliva self-test in this study, and for the SD Biosensor Ag-RDT conducted by trained staff in a previous study.^6^ In that study, sensitivities were 54.5% for oropharyngeal-nasal sampling and 68.4% for nasopharyngeal sampling in individuals with prior infection, and 75.8% and 75.0%, respectively, in individuals without prior infection. The low sensitivity in individuals with a prior infection may be explained by lower viral loads in this group (in the current study, 12/19 participants with a prior infection were below the viral load cut-off compared to 113/161 in those without a prior infection), with some of them potentially carrying viral RNA in the absence of a productive infection (i.e., no viral antigen production). Another explanation might be that individuals who have had COVID-19 have circulating anti-nuclear capsid (N) protein antibodies.^24^ These anti-N antibodies might bind to the N protein that is produced during the new infection, hampering the binding of monoclonal antibodies against the N-protein in the test device. It should be noted that we found a smaller reduced sensitivity of the BD Veritor Ag-RDT conducted by trained staff (oropharyngeal-nasal sampling) in individuals with and without a prior infection (64.6% versus 70.1%), so this effect may be test device-specific.^6^

### Strengths and limitations of this study

Strengths of this study include the large overall sample size covering multiple test sites nationwide, collection of samples for the reference and two Ag-RDTs in the same individuals within a few hours allowing for a head-to-head comparison of the two self-tests, sampling done by the participants themselves without any supervision conform the real-world context of self-testing, blinding of the index for the reference test result and vice versa, and the use of a proxy for infectiousness. Furthermore, the follow-up information showed that very few infections were missed by the molecular reference tests.

Our study also has some limitations. First, the reference standards that we used were molecular tests, but platforms and test kits used differed among the centralized laboratories. However, the diagnostic accuracies of all molecular tests used are similarly high^25,26^, and we therefore believe that this has not influenced our findings significantly. In addition, Ct values used to calculate viral loads were determined by different yet comparable platforms (supplementary material 3). Second, we used the viral load cut-off above which 95% of people with a positive RT-PCR test result had a positive virus culture as a proxy of infectiousness. Although this cut-off is not fully evidence based3, it is a best estimate based on current knowledge, and less arbitrary than using Ct cut-offs of 25 or 30 as is often done.^27,28^ In the current study, we relied on infectiousness viral load cut-offs that were determined in our previous study in a mainly unvaccinated population (the proportion of vaccinated individuals in the present study reached 85% at the end of the study), and when a different SARS-CoV-2 variant was dominant.^3^ Whether this would have impacted the applied viral load cut-offs is unknown, but vaccination itself did not influence any of the test sensitivities. Third, our sample size calculation was based on the primary analysis and the diagnostic accuracy parameters are less precise for the secondary stratified analyses. Fourth, participants were not blinded to the results of the saliva Ag-RDT when interpreting the result of the nasal Ag-RDT, which could have potentially biased the test outcome assessment of the nasal Ag-RDT. We do, however, believe that the impact of this limitation is small considering that the interpretation of the test results was considered (very) easy by >95% of participants, and the performance of each Ag-RDT was substantially different, and higher for the last performed nasal Ag-RDT. If outcome assessment was biased, the diagnostic performance of the self-tests would likely have been more similar. Fifth, we had some, though very limited, missing index test data (5%). We did not perform multiple imputation techniques because the group with missing data was very similar to the group with complete data, suggesting that data was missing completely at random.

### Policy implications

Ag-RDTs for self-use are widely available in the Netherlands. Until recently, the recommendation was to use them when asymptomatic prior to having contacts (such as going to school, events, or work), and visit a public health test site for molecular testing when symptomatic. Individuals whose self-test was positive are (still) asked to visit a public health test site for confirmatory testing. The SD Biosensor nasal self-test that we evaluated in this study is one of the self-tests that is commercially available in the Netherlands, although we do not know its market share. Our results indicate that the SD Biosensor nasal self-test sensitivity among individuals with mild symptoms is similar to Ag-RDT sensitivities found in studies where it was applied to professionally obtained upper respiratory tract samples. Based on those results, the Dutch Outbreak Management Team (OMT) that advises the Ministry of Health, Welfare, and Sports regarding COVID-19 policy recommended expanding nasal self-testing to individuals with mild symptoms. The OMT stressed that self-tests are not advised in vulnerable persons, in individuals meeting vulnerable persons, and in case of more severe symptoms, and that a negative self-test result is not sufficient for ending quarantine for contacts of a confirmed case. However, individuals testing negative by nasal self-testing would be allowed to go to work (if not working with vulnerable persons) or school despite their mild symptoms, preferably using mouth-nose masks and testing again a day later in case the initial test result was negative. Individuals testing positive by nasal self-testing would have to self-isolate and visit a public health test site for confirmatory molecular testing, to keep track of virus spreading and to allow for contact-tracing. All these nuances require careful communication, including on the implications of false-negative test results.

The very low sensitivity of the SD Biosensor nasal self-test in asymptomatic individuals is worrisome, even though the a-priori probability of being infected is lower in asymptomatic than symptomatic individuals. In addition, the potentially reduced sensitivity of the SD Biosensor nasal self-test in individuals who have had COVID-19 in the past is also worrisome. This is especially important because an increasing proportion of the population will have had COVID-19. We call for additional research in these two specific subgroups. We also recommend that persons who tested negative by a self-test continue to adhere to the general preventive measures such as physical distancing, wearing mouth-nose masks, and washing hands.

The SD Biosensor nasal self-test is only one of the commercially available self-tests. We recommend that all available self-tests are evaluated urgently by independent researchers, also addressing the relevant subgroups. Finally, in high-risk situations, such as testing of vulnerable people in care facilities, severely ill patients, or healthcare workers, we recommend molecular testing at all times, which is already in line with current policy.

## Supporting information

STARD Checklist

## Data Availability

All data produced in the present study are available upon reasonable request to the authors

## Acknowledgements

We thank the participants, and study staff at the participating public health service test sites, participating laboratories, the University Medical Center Utrecht, and RIVM for their contributions to the study. A special thanks to Esther Stiefelhagen, Renske Beekes, Sophie Neeleman, Roel Ensing, Wendy Mouthaan, Lieke Brouwer, and Timo Boelsums. Written permission was obtained from all five of them to list their names. ES, RB, SN, RE, WH, LB, and TB did not receive any compensation for their contributions.

## Contributors

KGMM initiated the study. ES, RPV, IKV, WvdB, SDP, EL, MH, RM, ZI, CW, IV, CRSN-I, WGHH, JAJWK, SvdH, JHHMvdW, and KGMM designed the study. ES, RPV, IKV, CRSN-I, and KGMM coordinated the study. WvdB, SDP, JS, RM, and ZI were responsible for laboratory analyses and data processing. ES, RPV, and IKV verified the underlying data. ES performed the statistical analysis in close collaboration with RPV and KGMM. ES, RPV, JHHMvdW and KGMM drafted the first version of the manuscript. All authors critically read the manuscript and provided feedback. All authors approved the submission of the current version of the manuscript. The corresponding author attests that all listed authors meet authorship criteria and that no others meeting the criteria have been omitted.

## Competing interests

All authors have completed the ICMJE uniform disclosure form at www.icmje.org/coi_disclosure.pdf and declare: support from the Dutch Ministry of Health, Welfare, and Sport for the submitted work; no financial relationships with any organisations that might have an interest in the submitted work in the previous three years; no other relationships or activities that could appear to have influenced the submitted work.

## Funding

This study was funded by the Dutch Ministry of Health, Welfare, and Sport. The funder had no role in the study design; collection, analysis, and interpretation of data; writing of the report; and decision to submit the paper for publication.

## Ethical approval

Not required because the study was judged by the METC Utrecht to be outside the scope of the Dutch Medical Research Involving Human Subjects Act (protocol No 21-470 /C). All participants signed an informed consent form before any study procedure.

## Data sharing

Individual participant data collected during the study will be available, after deidentification of all participants. Data will be available to researchers who provide a methodologically sound proposal to achieve the aims in the approved proposal. Proposals should be directed to the corresponding author to gain access to the data. Data requestors will need to sign a data sharing agreement.

The corresponding author (KGMM, the manuscript’s guarantor) affirms that the manuscript is an honest, accurate, and transparent account of the study being reported; that no important aspects of the study have been omitted; and that any discrepancies from the study as originally planned (and, if relevant, registered) have been explained.

## Additional information

The study protocol is available upon request by contacting Karel Moons at

## Notes

### Competing Interest Statement

The authors have declared no competing interest.

### Author Declarations

Ethical approval was not required because the study was judged by the METC Utrecht to be outside the scope of the Dutch Medical Research Involving Human Subjects Act (protocol No 21-470 /C). All participants signed an informed consent form before any study procedure.

