## Supplementary Material for "Accuracy and usability of saliva and nasal rapid antigen self-testing for detection of SARS-CoV-2 infection in the general population: a head-to-head comparison"

Table of contents

**Supplementary material 1: short questionnaire after performing both self-tests at home** 2

**Supplementary material 2: short questionnaire provided 10 days after test site visit** 5

**Supplementary material 3: Specimen collection, SARS-CoV-2 diagnostic testing, and SARS-CoV-2 virus culture procedures** 6

**Table S1 2x2 tables for primary and secondary analyses of the molecular reference test – saliva Ag-RDT comparison** 8

**Table S2 2x2 tables for primary and secondary analyses of the molecular reference test – nasal Ag-RDT comparison** 9

**Table S3 Follow-up information of participants that were tested again within 10 days after the initial test.** 10

**Figure S1 Sensitivities with 95% confidence intervals of the antigen rapid test-molecular reference standard test comparisons at viral load cut-off for infectiousness** 11

**Figure S2 Distribution of viral loads in participants with a positive molecular reference test, stratified by symptomatology at time of sampling.** 12

**Supplementary material 1: short questionnaire after performing both self-tests at home (translated from Dutch)**

1. What was the result of the saliva self-test?
   - ☐ Negative
   - ☐ Positive
   - ☐ I don't know: could not perform the test
   - ☐ I don't know: could not read the test result
   - ☐ Other, …………

2. What was the result of the nasal self-test?
   - ☐ Negative
   - ☐ Positive
   - ☐ I don't know: could not perform the test
   - ☐ I don't know: could not read the test result
   - ☐ Other, …………

3. What is the reason for testing?
   *Multiple answers possible*
   - ☐ I have (had) Covid-19 like symptoms
   - ☐ I am a close contact of a SARS-CoV-2 infected person
   - \- Date of last contact: ___ - ___ (day – month)
   
   I was notified because of:
   - \- The infected person is a household member □ yes / □ no
   - \- Received notification by CoronaMelder app (English: Corona notification app) □ yes / □ no
   - \- Received notification by public health service (by phone or letter) □ yes / □ no
   - \- Received notification by SARS-CoV-2 infected person □ yes / □ no
   - \- Other, ………
   - ☐ GP recommended a SARS-CoV-2 test
   - ☐ Travelled to a high-pandemic country
   - ☐ I performed a self-test and tested positive
   - \- What was the date of the positive self-test: ___ - ___ (day – month)
   - ☐ None of the above, but…….

4. Did you receive a Covid-19 vaccination?
   - ☐ No
   - ☐ Yes
   - \- If yes, which vaccine did you receive? ☐ Pfizer ☐ Moderna ☐ AstraZeneca ☐ Janssen ☐ Unknown
   - \- How many vaccinations did you receive? ☐ One ☐ Two
   - \- What was the date of the last vaccination? ___ - ___ - ___ (day – month – year)

5. Did you have had a positive SARS-CoV-2 test previously? If yes, how long ago?
   - ☐ No
   - ☐ Yes
   - \- If yes, how long ago? ☐ < 2 months ☐ 2-6 months ☐ 6-12 months ☐ >12 months

6. At this moment, do you have any COVID-19 like symptoms?
   - ☐ No ***GO TO QUESTION 9***
   - ☐ Yes

7. What COVID-19 like symptoms do you currently have?
   *Multiple answers possible*
   - ☐ Common cold
   - ☐ Shortness of breath
   - ☐ Fever
   - ☐ Coughing
   - ☐ Loss of taste or smell
   - ☐ Muscle ache
   - ☐ I have other symptoms: ……….

8. What was the moment you first experienced these symptoms?
   - ☐ Today
   - ☐ Yesterday
   - ☐ Two days ago
   - ☐ Three or more days ago

9. What is your highest level of education?
   - ☐ Primary
   - ☐ Secondary
   - ☐ College / University, Bachelor's Degree
   - ☐ College / University, Master's Degree

10. The saliva self-test instruction indicates you cannot eat, drink, smoke, brush your teeth or chew gum 30 minutes prior to the test, did you follow this instruction?
    - ☐ Yes
    - ☐ No
    - If no, I
      *Multiple answers possible*
      - ☐ Ate
      - ☐ drank
      - ☐ smoked
      - ☐ brushed my teeth
      - ☐ chewed gum

11. On a scale from 1 (very easy) to 5 (very hard), how difficult was it to perform the saliva self-test?

    Very easy 1 2 3 4 5 very hard

12. On a scale from 1 (very easy) to 5 (very hard), how difficult was it to read the result off the saliva self-test?

    Very easy 1 2 3 4 5 very hard

13. On a scale from 1 (very easy) to 5 (very hard), how difficult was it to perform the nasal self-test?

    Very easy 1 2 3 4 5 very hard

14. On a scale from 1 (very easy) to 5 (very hard), how difficult was it to read the result off the nasal self-test?

    Very easy 1 2 3 4 5 very hard

15. Do you agree with the following statements?

    The instruction for use of the saliva self-test was clear
    *Agree* *In doubt* *Disagree*

    I am confident I performed the saliva self-test correctly
    *Agree* *In doubt* *Disagree*

    I would use this saliva self-test at home if I could
    *Agree* *In doubt* *Disagree*

16. Do you prefer the saliva or nasal self-test?
    - ☐ No
    - ☐ Yes, I prefer the saliva self-test
    - ☐ Yes, I prefer the nasal self-test
    - If yes, why? ……….

17. Do you have suggestions for improvement of the instruction for use of the saliva self-test or other suggestions?
   ………

**Supplementary material 2: short questionnaire provided 10 days after test site visit (translated from Dutch)**

1. Did you develop any Covid-19 like symptoms since your last corona test 10 days ago (see next question for list of possible symptoms)?
   - ☐ No
   - ☐ Yes

2. What COVID-19 like symptoms do you currently have?
   *Multiple answers possible*
   - ☐ Common cold
   - ☐ Shortness of breath
   - ☐ Fever
   - ☐ Coughing
   - ☐ Loss of taste or smell
   - ☐ Muscle ache
   - ☐ I have other symptoms: ……….

3. What was the moment you first experienced these symptoms?
   - ☐ Today
   - ☐ Yesterday
   - ☐ Two days ago
   - ☐ Three or more days ago

4. Did you perform a corona test or was a corona test performed in the last 10 days since your corona test at the test site and the two self-tests performed for this study?
   - ☐ No ***THIS CONCLUDES THIS QUESTIONNAIRE***
   - ☐ Yes
   - If yes, what type of test was performed?
     - ☐ Test at a public health service test site
       - ☐ PCR
       - ☐ (Rapid) antigen test
       - ☐ I don't know
     - ☐ Test at a commercial test site
       - ☐ PCR
       - ☐ (Rapid) antigen test
       - ☐ I don't know
     - ☐ Self-test that I bought myself
     - ☐ Other, ………

5. What is the reason for testing?
   *Multiple answers possible*
   - ☐ I have (had) Covid-19 like symptoms
   - ☐ I am a close contact of a SARS-CoV-2 infected person
   - Date of last contact: ___ - ___ (day – month)

   I was notified because of:
   - The infected person is a household member □ yes / □ no
   - Received notification by CoronaMelder app (English: Corona notification app) □ yes / □ no
   - Received notification by public health service (by phone or letter) □ yes / □ no
   - Received notification by SARS-CoV-2 infected person □ yes / □ no
   - Other, ………

   - ☐ GP recommended a SARS-CoV-2 test
   - ☐ Travelled to a high-pandemic country
   - ☐ I performed a self-test and tested positive
   - What was the date of the positive self-test: ___ - ___ (day – month)
   - ☐ None of the above, but…….

6. What was the result of the corona test?
   - ☐ Positive (SARS-CoV-2 infection present)
   - ☐ Negative
   - ☐ No test result (test failed)

**Supplementary material 3: Specimen collection, SARS-CoV-2 diagnostic testing, and SARS-CoV-2 virus culture procedures**

**Specimen collection and SARS-CoV-2 diagnostic testing procedures**

*West Brabant region, Microvida laboratories at Bravis hospital in Roosendaal; samples from Roosendaal*

One superficial combined oropharyngeal and nasal swab (about 2.5 cm deep from the edge of the nostril) was taken per person at the Public Health Service test sites at the test sites in Roosendaal, placed in universal transport media (HiViralTM) with MagnaPure LC lysis- and binding buffer (Roche Diagnostics, The Netherlands), and transported to the Microvida laboratory in Roosendaal. RT-PCR was used as the reference standard test. RT-PCR was performed using the cobas SARS-CoV-2 test on the cobas 8800 platform (Roche Diagnostics International, Rotkreuz, Switzerland). Cycle threshold (Ct) values for the SARS-CoV-2 E-gene were converted to viral load (genome copies/ml) using an in-house established standard curve. The assay was used in accordance with the manufacturer's instructions; amplification curves and Ct values were interpreted using the manufacturer's interpretation algorithms, which complied with the European in-vitro diagnostic devices directive.

*Central- and Northeast Brabant region, Microvida laboratories at Elisabeth Twee Steden Ziekenhuis (ETZ) in Tilburg; samples from Tilburg*

One deep combined oropharyngeal-nasopharyngeal swab (>5 cm deep from the edge of the nostril) was taken per person at the Public Health Service test site in Tilburg, placed in 1x Tris-EDTA buffer (Sigma-Aldrich), and transported to the Microvida laboratory in Tilburg. One of two molecular tests were used as the reference test: the SARS-CoV-2 assay on the Alinity M molecular platform (Abbott) according to manufacturer's instructions, or an in-house RT-PCR as previously described using automated DNA extraction on a Qiagen Symphony with subsequent RT-PCR on a Qiagen Rotor-Gene platform.[1] RT-PCR positive samples were further analyzed on a cobas 8800 platform (in the Microvida laboratory in Roosendaal) to obtain Ct values for viral load calculation. Samples were stored at -80°C within 24 hours and thawed once for the cobas 8800 analysis.[2] In case of discrepant results (Alinity/in-house positive, cobas 8800 negative), a Ct value of 40 was assigned for subsequent analysis.

*Rotterdam city region, Erasmus Medical Center Viroscience laboratory: samples from south of Rotterdam (Zuidland)*

One deep combined OP-N swab (>5 cm deep from the edge of the nostril) swab was taken per person at the Public Health service test site in the Rotterdam-Rijnmond region (Zuidland), placed directly in universal transport media (HiViralTM), and transported to the Erasmus MC Viroscience diagnostic laboratory. RT-PCR was used as the reference standard test. RT-PCR testing was performed in virus transport medium using the cobas SARS-CoV-2 test on the cobas 6800 platform. The assay was used in accordance with the manufacturer's instructions; amplification curves and cycle threshold values were interpreted using the manufacturer's interpretation algorithms, which complied with the European in-vitro diagnostic devices directive.

**Viral load calculation**

We used a viral load cut-off as a proxy of infectiousness of ≥5.2 log10 SARS-CoV-2 E-gene copies/mL, which was the viral load cut-off above which 95% of people with a positive molecular test had a positive virus culture in a recent previous study by our group.[1] For that study, the Erasmus MC Viroscience laboratory created a standard curve by testing dilutions of a specific quantified E-gene transcript (primary standard) available from the European Virus Archive (EVAg[4]) with the RT-PCR described by Corman et al.[3] The relation between Ct value and number of E-gene copies/ml was determined by linear regression analysis. Based on this calibration curve a secondary standard derived from cell-cultured virus was prepared and quantified. Serial dilutions of this secondary standard were tested by RT-PCR to prepare a secondary standard curve by linear regression.

In that previous study, we determined whether the cobas PCR platform at the Microvida laboratory provided comparable data to the cobas PCR platform at the Erasmus Medical Center Viroscience laboratory by having both laboratories test the same SARS-CoV-2 viral load panel obtained from the National Public Health Institute (RIVM). The Ct values generated in the two laboratories corresponded well. Therefore, we concluded that the same formula to convert Ct values into viral loads (copies/ml) could be used for the West-Brabant and Rotterdam laboratories This formula was $62.5 * e^{\frac{43.1-Ct}{1.607}}$. The Microvida laboratory used different medium/reagent volumes per swab depending on the location. For each, a factor $X$ was calculated based on the swab volume, dilution, and total sample volume, to allow alignment across samples with respect to the viral load in the Rotterdam standard. For example, if the sample volume is equal (3 ml in Tilburg compared to 3 ml in Rotterdam), but the dilution factor is higher, e.g., by the addition of lysis buffer (3 in Tilburg compared to 2 in Rotterdam), the effective viral sample would be lower than in the Rotterdam standard, thereby underestimating the actual viral load when using the above formula. To account for this, a factor $X < 1$ was used, here 6/9. If dilution was lower, a factor $X > 1$ was used. The conversion formula used therefore was $62.5 * e^{\frac{43.1-Ct}{1.607}} / X$.

**Self-tests**

*Saliva Ag-RDT*

The saliva Ag-RDT used in this study was the COVID-19 Antigen Rapid Test (Oral Fluid) for Self-testing by Hangzhou AllTest Biotech Co., Ltd., a CE-marked Ag-RDT. According to the instructions for use, participants had to cough deeply 3 to 5 times after which saliva could be spit in the test tube. After addition of buffer fluid, and squeezing the test tube 10-15 times, two droplets of the solvent could be added to the testing cassette. After 15 minutes, but no later than 20 minutes, the participant could read the test result visually from the testing cassette.

*Nasal Ag-RDT*

The nasal Ag-RDT used in this study was the SD Biosensor SARS-CoV-2 Rapid Antigen Self-Test Nasal, distributed by Roche Diagnostics, a CE-marked Ag-RDT. According to the instructions for use, a sample should be taken by placing a nasal swab 2 cm deep in the nose and twisting it 4 times for 15 seconds, once in each nostril. Afterwards the nose swab had to be placed in a test tube that was filled by buffer fluid and twisted 10 times during which the test tube had to be squeezed lightly. While squeezing the tube, the swab could be removed from the tube, while leaving all the test fluid in the tube. Then, 4 droplets of the solvent had to be added to the testing cassette. After 15 minutes, but no later than 30 minutes, the participant had to read the test result visually from the testing cassette.

*Analytical performance*

The analytical performances of the Hangzhou AllTest Biotech and the SD Biosensor lateral flow test devices were assessed using a national External Validation Quality Assessment panel (LEQA) for SARS-CoV-2 Ag-RDTs. The LEQA Ag panel contained four different SARS-CoV-2 strains, including:
hCoV-19/Netherlands/NoordBrabant_10003/2020 (B.11, 19A);
hCoV-19/Netherlands/GE-RIVM-20300/2020 (B.1.177, 20AEU1);
hCoV-19/Netherlands/NH-RIVM-20432/2020 (B.1.1.7, 20B/501Y.V1); and
hCoV-19/Netherlands/NB-RIVM-20274/2020 (B.1.5, 20A).
These strains had been deactivated by 2-hour incubation with Triton X-100 at room temperature.
The results of the panel assessment are presented in the Table below and show that both devices had good analytical performance. Furthermore, the analytical performance of the two devices were very similar.

| LEQA Ag panel | SARS-CoV-2 strain | Concentration TCID50/ml | Hangzhou AllTest Biotech | SD Biosensor |
|---|---|---|---|---|
| LEQAA1_Ag21-01 | B.1.177 20A.EU1 | 1334 | ++ | ++ |
| LEQAA1_Ag21-02 | B.11; 19A; wildtype | 7499 | ++ | ++ |
| LEQAA1_Ag21-03 | B.1.1.7 20B/501Y.V1 | 23714 | ++ | ++ |
| LEQAA1_Ag21-04 | B.11; 19A; wildtype | 75 | + | (+) |
| LEQAA1_Ag21-05 | B.1.1.7 20B/501Y.V1 | 237 | + | + |
| LEQAA1_Ag21-06 | B.1.177 20A.EU1 | 13335 | ++ | ++ |
| LEQAA1_Ag21-07 | B.1.5 20A | 2371 | ++ | ++ |
| LEQAA1_Ag21-08 | B.1.177 20A.EU1 | 133 | ++ | + |
| LEQAA1_Ag21-09 | Negative | | - | - |
| LEQAA1_Ag21-10 | B.1.1.7 20B/501Y.V1 | 2371 | ++ | ++ |
| LEQAA1_Ag21-11 | B.11; 19A; wildtype | 7 | (+) | - |
| LEQAA1_Ag21-12 | B.1.5 20A | 23714 | ++ | ++ |
| LEQAA1_Ag21-13 | B.11; 19A; wildtype | 750 | ++ | + |
| LEQAA1_Ag21-14 | B.1.5 20A | 237 | ++ | + |
| LEQAA1_Ag21-15 | B.11; 19A; wildtype | 74989 | ++ | ++ |

TCID50 = Median Tissue Culture Infectious Dose
- = negative; (+) = faint positive; + = positive; ++ = strong positive

**Table S1 2x2 tables for primary and secondary analyses of the molecular reference test – saliva Ag-RDT comparison**

| Analysis | Subgroup | | Ref+ | Ref- | Total |
|---|---|---|---|---|---|
| **Primary** | | | **Ref+** | **Ref-** | **Total** |
| | | **Test+** | **85** | **26** | **111** |
| | | **Test-** | **97** | **2595** | **2692** |
| | | **Total** | **182** | **2621** | **2803** |
| **Secondary (stratified):** | | | | | |
| Infectiousness viral load cut-off¶ | | | Ref+ | Ref- | Total |
| | | Test+ | 78 | 32 | 110 |
| | | Test- | 64 | 2614 | 2678 |
| | | Total | 142 | 2646 | 2788 |
| Symptoms present at sampling‡: | Yes | | Ref+ | Ref- | Total |
| | | Test+ | 77 | 25 | 102 |
| | | Test- | 74 | 2130 | 2204 |
| | | Total | 151 | 2155 | 2306 |
| | No | | Ref+ | Ref- | Total |
| | | Test+ | 8 | 1 | 9 |
| | | Test- | 23 | 452 | 475 |
| | | Total | 31 | 453 | 484 |
| Vaccinated (at least 1 one)[@]: | Yes | | Ref+ | Ref- | Total |
| | | Test+ | 54 | 22 | 76 |
| | | Test- | 61 | 2253 | 2314 |
| | | Total | 115 | 2275 | 2390 |
| | No | | Ref+ | Ref- | Total |
| | | Test+ | 31 | 4 | 35 |
| | | Test- | 36 | 333 | 369 |
| | | Total | 67 | 337 | 404 |
| Previous SARS-CoV-2 infection[§]: | Yes | | Ref+ | Ref- | Total |
| | | Test+ | 4 | 3 | 7 |
| | | Test- | 16 | 343 | 359 |
| | | Total | 20 | 346 | 366 |
| | No | | Ref+ | Ref- | Total |
| | | Test+ | 81 | 23 | 104 |
| | | Test- | 81 | 2242 | 2323 |
| | | Total | 162 | 2265 | 2427 |
| Sex[†]: | Female | | Ref+ | Ref- | Total |
| | | Test+ | 41 | 13 | 54 |
| | | Test- | 67 | 1590 | 1657 |
| | | Total | 108 | 1603 | 1711 |
| | Male | | Ref+ | Ref- | Total |
| | | Test+ | 44 | 13 | 57 |
| | | Test- | 29 | 995 | 1024 |
| | | Total | 73 | 1008 | 1081 |
| Age [years] [#]: | ≥16 to ≤40 | | Ref+ | Ref- | Total |
| | | Test+ | 38 | 15 | 53 |
| | | Test- | 54 | 1343 | 1397 |
| | | Total | 92 | 1358 | 1450 |
| | >40 to ≤65 | | Ref+ | Ref- | Total |
| | | Test+ | 36 | 9 | 45 |
| | | Test- | 36 | 1054 | 1090 |
| | | Total | 72 | 1063 | 1135 |
| | >65 | | Ref+ | Ref- | Total |
| | | Test+ | 11 | 2 | 13 |
| | | Test- | 6 | 189 | 195 |
| | | Total | 17 | 191 | 208 |

¶Viral load cut-off for infectiousness, defined as viral load above which 95% of people with a positive RT-PCR test result had a positive viral culture[6], was 5.2 log10 SARS-CoV-2 E-gene copies/mL. Viral load unavailable for 19 individuals.
‡Symptoms not available for 13 participants.
[@] COVID-19 vaccination status not available for 9 participants.
[§] Previous SARS-CoV-2 infection information not available for 10 participants.
[†] Sex not available for 11 participants.
[#] Age not available for 10 participants.

**Table S2 2x2 tables for primary and secondary analyses of the molecular reference test – nasal Ag-RDT comparison**

| Analysis | Subgroup | | | | |
|---|---|---|---|---|---|
| **Primary** | | | **Ref+** | **Ref-** | **Total** |
| | | **Test+** | **124** | **12** | **136** |
| | | **Test-** | **56** | **2627** | **2683** |
| | | **Total** | **180** | **2639** | **2819** |
| **Secondary (stratified):** | | | | | |
| Infectiousness viral load cut-off¶ | | | Ref+ | Ref- | Total |
| | | Test+ | 120 | 13 | 133 |
| | | Test- | 23 | 2648 | 2671 |
| | | Total | 143 | 2661 | 2804 |
| Symptoms present at sampling‡: | Yes | | Ref+ | Ref- | Total |
| | | Test+ | 117 | 10 | 127 |
| | | Test- | 32 | 2160 | 2192 |
| | | Total | 149 | 2170 | 2319 |
| | No | | Ref+ | Ref- | Total |
| | | Test+ | 7 | 2 | 9 |
| | | Test- | 24 | 454 | 478 |
| | | Total | 31 | 456 | 487 |
| Vaccinated (at least 1 one)[@]: | Yes | | Ref+ | Ref- | Total |
| | | Test+ | 75 | 10 | 85 |
| | | Test- | 32 | 2272 | 2304 |
| | | Total | 107 | 2282 | 2389 |
| | No | | Ref+ | Ref- | Total |
| | | Test+ | 49 | 2 | 51 |
| | | Test- | 24 | 346 | 370 |
| | | Total | 73 | 348 | 421 |
| Previous SARS-CoV-2 infection[§]: | Yes | | Ref+ | Ref- | Total |
| | | Test+ | 7 | 3 | 10 |
| | | Test- | 12 | 350 | 362 |
| | | Total | 19 | 353 | 372 |
| | No | | Ref+ | Ref- | Total |
| | | Test+ | 117 | 9 | 126 |
| | | Test- | 44 | 2267 | 2311 |
| | | Total | 161 | 2276 | 2437 |
| Sex[†]: | Female | | Ref+ | Ref- | Total |
| | | Test+ | 73 | 7 | 80 |
| | | Test- | 33 | 1604 | 1637 |
| | | Total | 106 | 1611 | 1717 |
| | Male | | Ref+ | Ref- | Total |
| | | Test+ | 51 | 5 | 56 |
| | | Test- | 22 | 1012 | 1034 |
| | | Total | 73 | 1017 | 1090 |
| Age [years] [#]: | ≥16 to ≤40 | | Ref+ | Ref- | Total |
| | | Test+ | 62 | 7 | 69 |
| | | Test- | 31 | 1363 | 1394 |
| | | Total | 93 | 1370 | 1463 |
| | >40 to ≤65 | | Ref+ | Ref- | Total |
| | | Test+ | 51 | 4 | 55 |
| | | Test- | 19 | 1065 | 1084 |
| | | Total | 70 | 1069 | 1139 |
| | >65 | | Ref+ | Ref- | Total |
| | | Test+ | 11 | 1 | 12 |
| | | Test- | 5 | 189 | 194 |
| | | Total | 16 | 190 | 206 |

¶Viral load cut-off for infectiousness, defined as viral load above which 95% of people with a positive RT-PCR test result had a positive viral culture[6], was 5.2 log10 SARS-CoV-2 E-gene copies/mL. Viral load unavailable for 19 individuals.
‡Symptoms not available for 13 participants.
[@] COVID-19 vaccination status not available for 9 participants.
[§] Previous SARS-CoV-2 infection information not available for 10 participants.
[†] Sex not available for 12 participants.
[#] Age not available for 11 participants.

**Table S3 Follow-up information of participants that were tested again within 10 days after the initial test.** Information was not available for 794 (28.3%) and 796 (28.2%) of participants with saliva and nasal Ag-RDT result available, respectively. Of the remaining participants 1758 (87.5%) and 1772 (87.6%) were not tested within 10 days after the initial test.

| Test result available from the reference test and… | Saliva Ag-RDT | Nasal Ag-RDT |
|---|---|---|
| Sample size (Test during 10-day follow-up) | N = 251/2009 (12.5%) | N = 251/2029 (12.4%) |
| Testing information, n (%)[!] | | |
| Symptomatic | 74 (29.5) | 74 (29.5) |
| Asymptomatic – close contact of confirmed SARS-CoV-2 infected household member | 0 (0.0) | 0 (0.0) |
| Asymptomatic – close contact of other confirmed SARS-CoV-2 infected individual | 40 (15.9) | 41 (16.3) |
| Asymptomatic – other | 137 (54.6) | 136 (54.2) |
| Type of test, n (%) | | |
| public health service COVID-19 test sites | 71 (28.3) | 70 (27.9) |
| RT-PCR test | 44 (62.0) | 44 (62.9) |
| Ag-RDT | 2 (2.8) | 2 (2.9) |
| Unknown | 25 (35.2) | 24 (34.2) |
| Commercial test site | 24 (9.6) | 24 (9.6) |
| RT-PCR test | 5 (20.8) | 5 (20.8) |
| Ag-RDT | 16 (66.7) | 16 (66.7) |
| Unknown | 3 (12.5) | 3 (12.5) |
| Self-test | 126 (50.2) | 127 (50.6) |
| Other (e.g., via work, school) | 30 (12.0) | 30 (12.0) |
| Test result | | |
| Positive | 8 (3.2) | 8 (3.2) |
| Negative | 240 (95.6) | 240 (95.6) |
| Unknown | 3 (1.2) | 3 (1.2) |

**Figure S1** Sensitivities with 95% confidence intervals of the antigen rapid test-molecular reference standard test comparisons at viral load cut-off for infectiousness (i.e., viral load <5.2 log10 SARS-CoV-2 E-gene copies/mL is considered as a negative reference test result), stratified according to symptomatology, COVID-19 vaccination status, previous infection status, sex, and age. The vertical line indicates the sensitivity of the Ag-RDT in the overall study population before application of a viral load cut-off (see Figure 2), and the number of positive molecular tests out of the total or subgroup between parentheses.

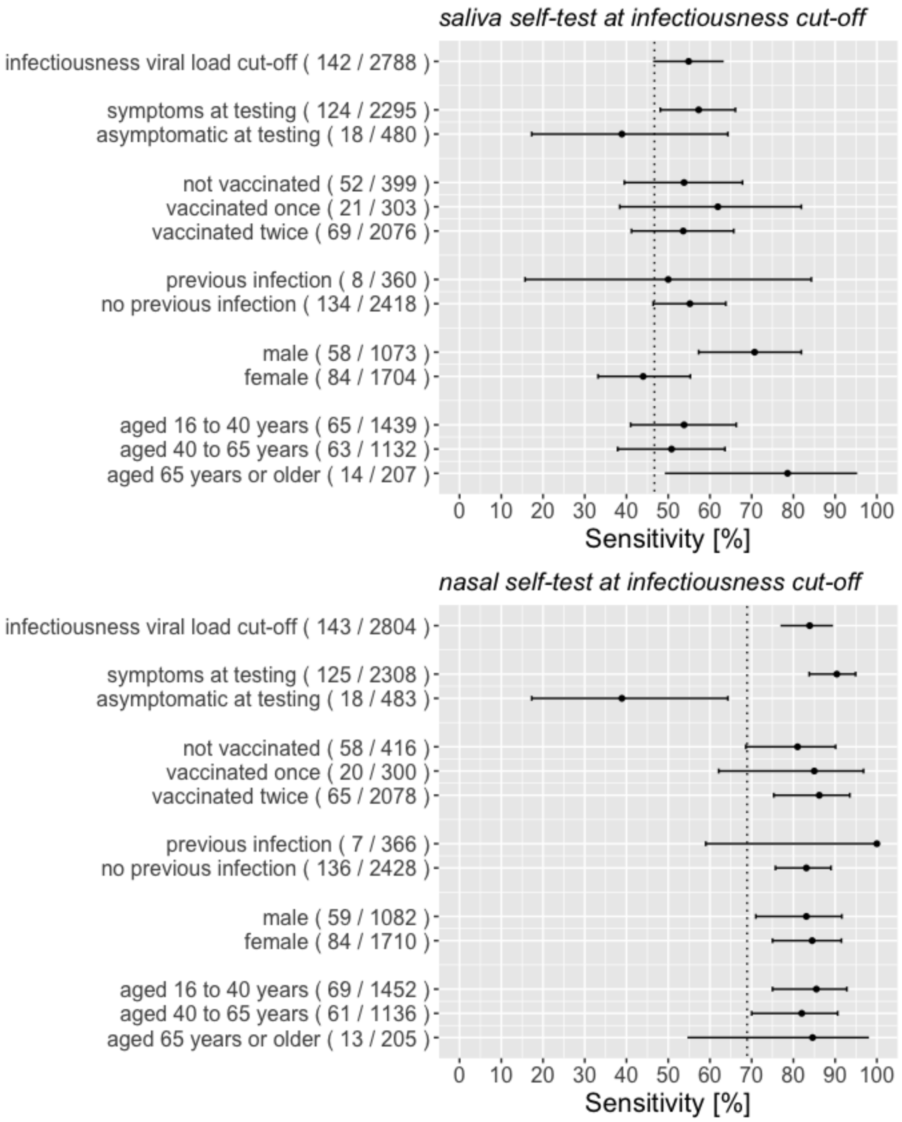

**Figure S2** Distribution of viral loads in participants with a positive molecular reference test, stratified by symptomatology at time of sampling.

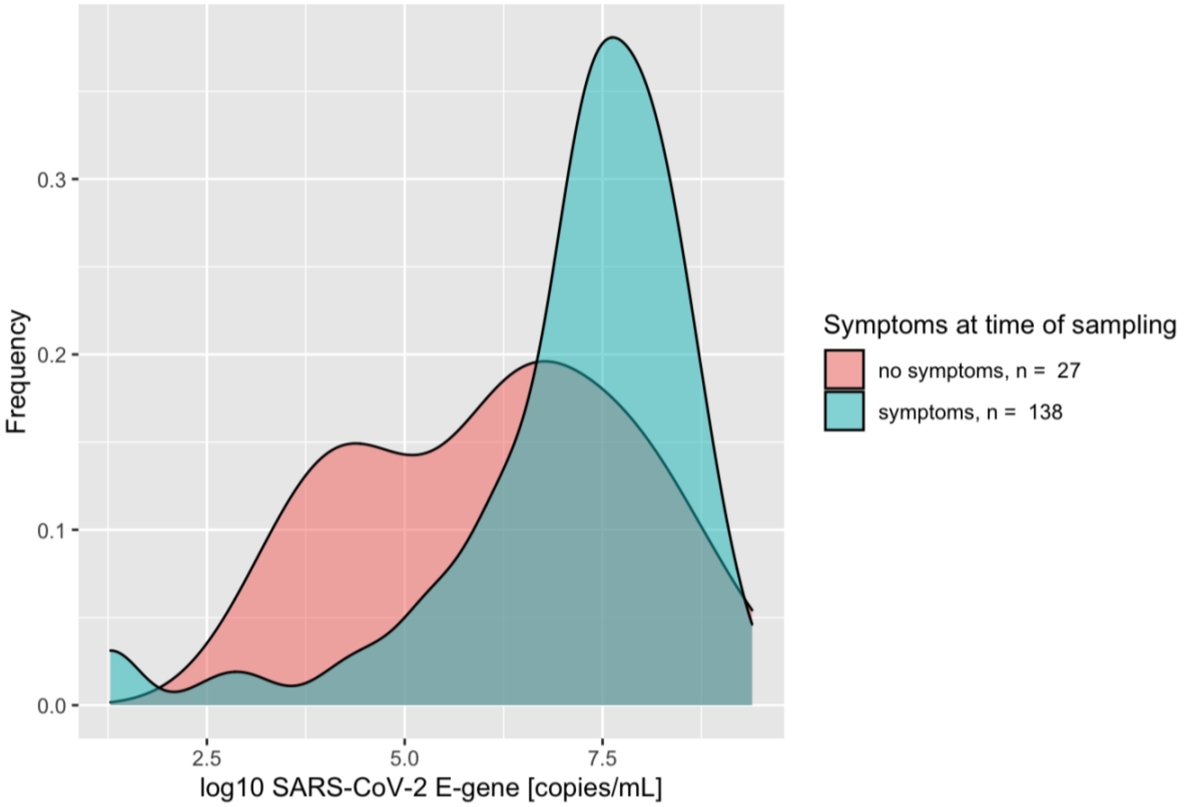
